# Association between Socioeconomic Status and Cardiovascular Disease by sex: Mediating roles of health behavior, depression, and unmet medical needs

**DOI:** 10.1101/2025.05.01.25326837

**Authors:** Jiwon Choi, Sung-il Cho

## Abstract

**Introduction:** Cardiovascular disease (CVD) is a leading cause of death, and numerous studies have examined its risk factors. Low socioeconomic status (SES) increases the risk of CVD and contributes to disparities in health outcomes through the unequal distribution of intermediary factors. Although health behaviors, psychosocial factors, and access to healthcare are unequally distributed across socioeconomic groups, the mechanisms remain unclear. Therefore, this study aims to identify the pathways between SES and CVD.

**Methods:** Data from the Korea Health Panel Survey from 2009 to 2018 were utilized. A total of 11,397 participants aged 30 and above, with no prior diagnosis of CVD and no missing responses, were included in the study. The exposure variable was SES, derived from latent class analysis, while the outcome was CVD incidence. The mediators included health behavior, depression symptoms, and unmet medical needs. The causal mediation analysis with the accelerated failure time (AFT) model was applied to assess the indirect effects.

**Results:** Three latent classes of SES—low, medium, and high—were derived using four variables: income, education, working status, and health insurance. A significant direct effect was observed between low SES and increased CVD risk in women compared to those with medium SES. Additionally, depression indirectly reduced the average survival time from CVD by 5.9% among women.

**Conclusions:** Indirect effects of depression among women were observed in the SES-CVD pathway. These indirect pathways help clarify the mechanisms behind social disparities in health outcomes and highlight the need for sex-stratified policy approaches to prevent CVD.

## 1. Introduction

Cardiovascular disease (CVD) is a leading cause of death worldwide. In 2020, CVD was responsible for approximately 19.05 million deaths globally ^1^. In South Korea, cardiovascular disease (CVD) was the leading cause of death until 1999 and has since remained the second most common cause, continuing to be a major contributor to mortality ^2^. Also, the years of life lost (YLLs) due to circulatory system diseases are estimated at 824 per 100,000 population ^3^. Additionally, the disability-adjusted life-year (DALYs) at 3,226 per 100,000 population shows a continuous increase over time, suggesting a steady rise in the overall CVD burden in Korea.

CVD incidence and mortality rates are affected by many cardiovascular risk factors (CVRFs), including smoking, alcohol consumption, lack of physical activity, obesity, and diabetes ^4^. Social, environmental, and economic factors also influence the incidence of CVD, and socioeconomic status (SES) has been identified as a significant predictor of CVD and its associated risk factors ^5,6^. Individuals with low SES exhibit higher CVD incidence and mortality rates, likely due to the higher prevalence of CVRFs and having more difficulty in accessing treatment and healthcare services ^7^.

In epidemiological studies, SES is assessed based on factors such as education, income, and occupation or employment status, which is also confirmed in CVD research ^5,8,9^. However, relying on a single element of SES fails to capture the variation in SES and could obscure significant social gradients in health ^10,11^. Therefore, several researchers have suggested using multiple variables as indicators of SES ^12,13^. However, including more than one SES indicator in regression models may violate the collinearity assumptions due to the high correlation ^14^. Composite indices incorporating multiple SES indicators are being developed to provide more comprehensive aspects to address this issue.

It is important to explore how to combine multiple measures of SES in a meaningful way, and latent class analysis (LCA) has emerged as a useful method. LCA is a person-centered approach that categorizes a population into mutually exclusive groups based on observable indicator variables ^15,16^. LCA models enable the meaningful classification of SES based on relevant indicators, facilitating the characterization of SES ^14^. Research using LCA to select proxy variables for composite aspects of SES is increasing ^17–19^. To address the limitations of existing single indicators of SES, our study was conducted to apply LCA using representative SES measures.

The social determinants of health (SDoH) — including social, environmental, economic, and psychosocial factors — play a critical role in the incidence of CVD and its associated risk factors ^20^. The Commission on Social Determinants of Health (CSDH) proposed a conceptual framework in which social contexts generate stratification, assigning individuals to different social positions ^21^. These positions influence health outcomes through material, psychosocial, behavioral/biological factors, and the health system. Health inequality arises when those intermediary factors are unevenly distributed among different socioeconomic classes. A better understanding of these mechanisms can inform public health interventions by identifying modifiable risk factors that mediate or interact with the effects of SES ^22^.

The potential pathways through which CVD is influenced can be explained through physiologic, health behaviors, environmental factors, and the healthcare system ^20,23^. Economic stability includes factors influencing income and employment status, affecting CVD through various direct and indirect pathways ^23^. The mechanism between economic stability and CVD can be explained by the pathways such as physiologic (e.g., depression, chronic stress) and health behaviors (e.g., smoking, physical inactivity, unhealthy diet) factors ^24–26^. Educational attainment may be associated with CVD risk through physiological, behavioral, and environmental factors as intermediaries ^27^. Additionally, income level and employment status can influence health insurance coverage and access to medical care, ultimately acting as intermediary factors that affect cardiovascular health ^28^.

The mediators were selected based on the intermediary determinants of the framework from the CSDH, including smoking status, physical activity, depression symptoms, and unmet medical needs. Socioeconomic disadvantage is linked to unhealthy behaviors like smoking and physical inactivity, which are subsequently linked to an elevated risk of CVD ^23,25^. There have been studies that SES might increase the risk of CVD through its relationship with depression by disrupting stress-response pathways such as the sympathetic nervous system ^29^. The health system becomes particularly relevant through the issue of access, and unmet medical needs serve as a proxy indicator of the lack of access to healthcare ^21,30^. Income level and employment status can influence access to medical care, affecting cardiovascular health in various ways ^23,28^.

Despite previous research, important gaps remain. First, prior studies usually use single variables (e.g., income, occupation, education) to represent the individual level of SES, which only partly reflects its multiple dimensions of SES ^8^. However, it is critical to construct a comprehensive variable that comprises varying aspects of SES ^12^. Second, there is a lack of studies that explore the pathways of intermediary factors within the framework of SDoH. Broader inequities in care and risk factors remain underexplored within a comprehensive SDoH framework ^23^. Therefore, our study aimed to examine the pathway between SES and CVD by quantifying the extent to which the effect of SES on the risk of CVD is mediated through intermediary factors (i.e., behavioral factors, depression, and unmet medical needs) based on the SDoH framework.

## 2. Methods

### 2.1. Data and study population

This study utilized the Korea Health Panel Survey (KHPS) data (version 1.7.3.), which was conducted annually from 2008 to 2018 by the Korea Institute for Health and Social Affairs (KIHASA) and the National Health Insurance Service (NHIS) consortium. This panel uses a multi-stage stratified probability sampling method based on samples extracted from the 90% complete enumeration data of the 2005 Population and Housing Census across 16 districts nationwide. The original panel was initiated in 2008 with approximately 8,000 households, and approximately 2,500 households were added in 2014 to compensate for attrition. KHPS provides disease, healthcare utilization, medical expenditure, and health-related behaviors. Since information on health behaviors (e.g., smoking habits and regular exercise) was collected during the second phase of the survey in 2009, this study utilized data from that year onward. The raw data are accessible to everyone and can be obtained by submitting a requisition form through the official KHPS website (www.khp.re.kr:444/).

From 20,395 individuals with at least one recorded entry between 2009 and 2013, a moving index was applied during the entry period to account for each individual’s enrollment date. We selected the study population through the following exclusions: 430 individuals with a history of CVD before the index date, 7,335 individuals who were under 30 years old at baseline, and 1,233 individuals for whom additional surveys on health behaviors, income, and depression symptoms were either not conducted or not responded to (**Figure 1**). Consequently, a total of 11,397 participants were included in the study.

**Figure 1.**
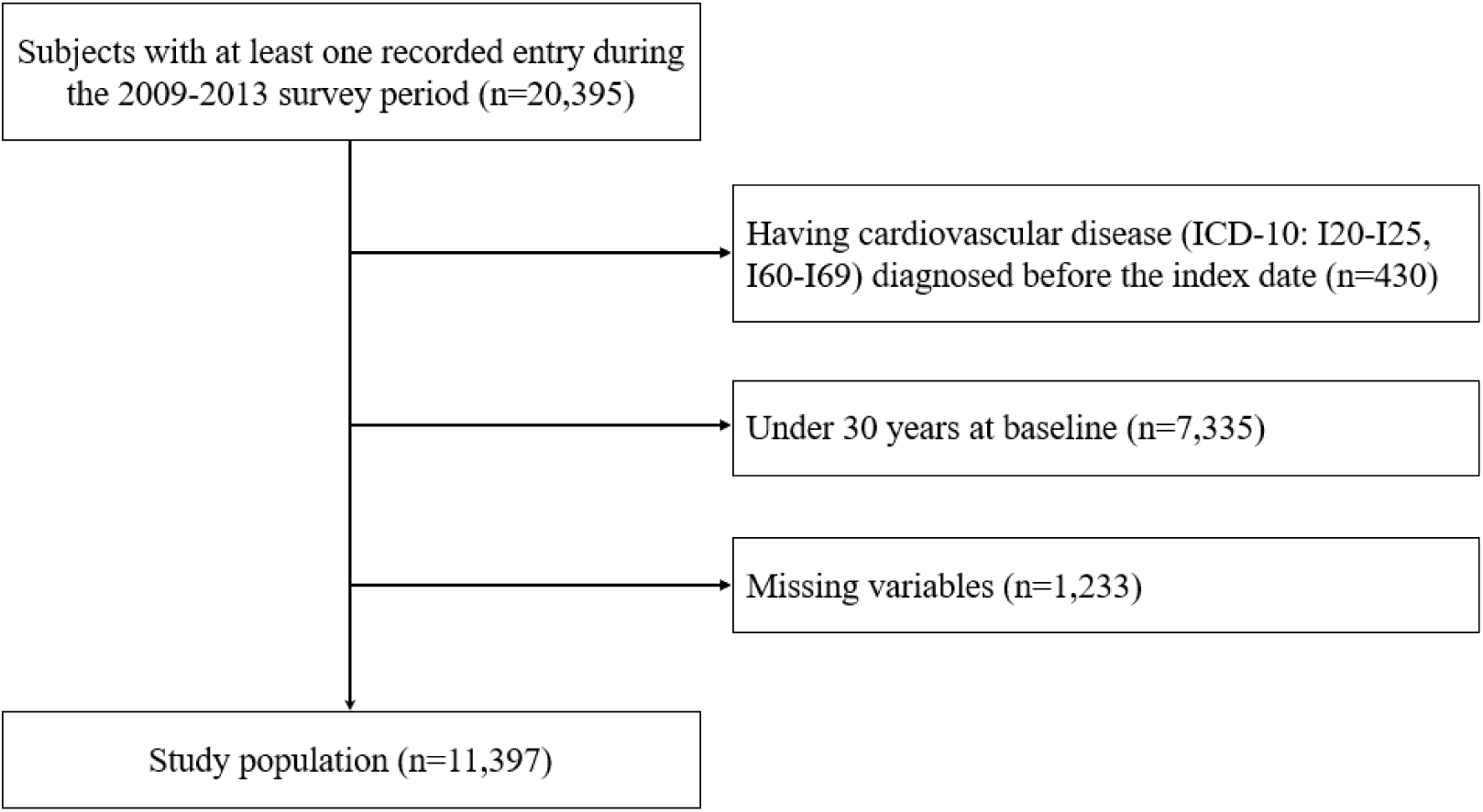
Selection of study population Abbreviation: ICD-10, International Classification of Diseases 10th edition.

### 2.2. Variables

The exposure variable is SES, defined by latent classes using four variables: self-reported household income, education level, working status, and health insurance. Household income was calculated by summing the labor income of all household members and capital income for the past year, then equivalized by taking the square root of the number of household members. It was classified into three groups by quintiles, with the 1st quintile categorized as low, the 2nd to 4th quintiles as middle, and the 5th quintile as high ^31^. Education level was classified into three categories: below elementary school graduation, middle and high school graduates, and those with a college degree or higher. Working status was classified based on the question, “What is your employment status?” Those who were not wage workers were classified as “none”, regular employees were classified as “full-time”, and other categories, such as temporary and daily workers, were classified as “part-time”. Health insurance was classified based on the type of coverage: those with national merit special benefits or foreign nationals not enrolled were classified as “none”, those eligible for medical aid (1st and 2nd types) were classified as “medical aid”, and those enrolled in health insurance were classified as “NHIS”. SES was derived through LCA by considering three levels for the following four variables. Consequently, three latent classes were identified, which can be interpreted as high, medium, and low SES.

The main outcome was newly diagnosed CVD cases defined by the KHPS’s own disease classification and the International Classification of Diseases 10th edition (ICD-10). Following the American Heart Association guidelines, CVD is defined as ischemic heart disease (ICD-10: I20–I25) or stroke (ICD-10: I60–I69) ^32–34^. The participants were censored by a CVD event, death, or 31 December 2018, whichever occurred first.

We hypothesized potential mediators for the causal pathway between SES and incidence of CVD, as shown in **Figure 2**. Smoking status was categorized into three groups: never-smoker, former smoker, and current smoker. Moderate physical activity was assessed by asking participants “how many days in the past week they engaged in at least 10 minutes of activity that caused them to breathe slightly harder and experience an increase in heart rate”, and the responses were coded as 0, 1-2, 3-5, or 6-7 days. In this study, participants who responded “yes” to the question, “Have you felt so sad or unhappy that it interfered with your daily life for more than two weeks in the past year?” were defined as experiencing depression symptoms. Unmet medical needs were assessed using the question, “In the past year, have you ever needed treatment or tests but did not receive them?” Individuals who responded affirmatively regarding their needs were defined as having unmet medical needs.

**Figure 2.**
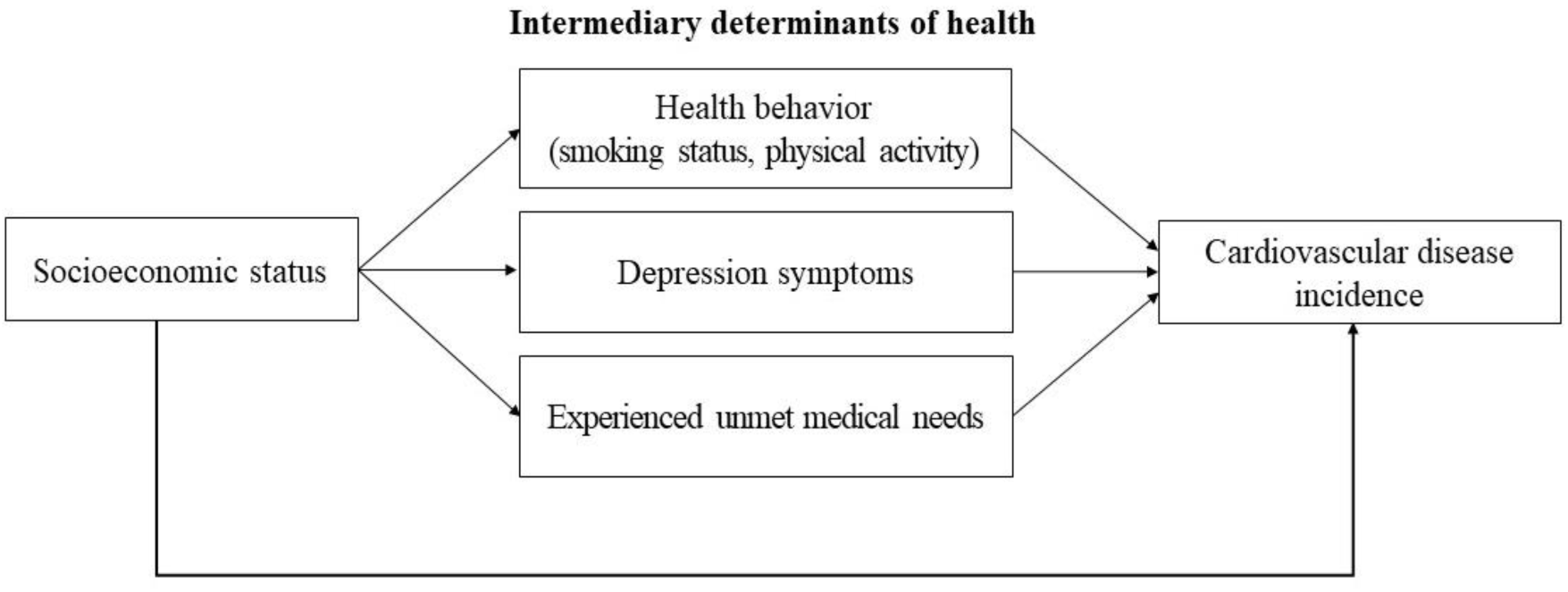
The direct and indirect pathways between socioeconomic status and cardiovascular disease

The covariates were defined as age, sex, residence, and marital status, based on previous studies ^17,35–37^. Age was categorized into 30-39, 40-49, 50-59, and 60 and above, while residence was classified as urban for Seoul and Gyeonggi and rural for other areas. Marital status was categorized as never married, married, separated/divorced, and widowed. The variables for the demographic characteristics of the study population are presented in **Table 1**.

**Table 1.** Baseline characteristics of the study population (n = 11,397)

| **Variables** | **N (%)** |
| --- | --- |
| **Sex** |  |
| Men | 5,126 (44.98) |
| Women | 6,271 (55.02) |
| **Age** |  |
| 30-39 | 2,698 (23.67) |
| 40-49 | 2,850 (25.01) |
| 50-59 | 2,304 (20.22) |
| 60+ | 3,545 (31.10) |
| **Residence** |  |
| Urban | 2,349 (20.61) |
| Rural | 9,048 (79.39) |
| **Marital status** |  |
| Never married | 802 (7.04) |
| Married | 9,123 (80.05) |
| Separated/Divorced | 396 (3.47) |
| Widowed | 1,076 (9.44) |
| **Household income** |  |
| Low (1<sup>st</sup> quintile) | 1,840 (16.14) |
| Middle (2<sup>nd</sup>-4<sup>th</sup> quintiles) | 7,062 (61.96) |
| High (5<sup>th</sup> quintiles) | 2,495 (21.89) |
| **Education level** |  |
| ≤ Elementary school | 2,674 (23.46) |
| Middle-high school | 5,362 (47.05) |
| ≥ College | 3,361 (29.49) |
| **Working status** |  |
| None | 7,017 (61.57) |
| Part-time | 3,251 (28.53) |
| Full-time | 1,129 (9.91) |
| **Health insurance** |  |
| None | 13 (0.11) |
| Medical aid | 437 (3.83) |
| NHIS | 10,947 (96.05) |
| **Smoking status** |  |
| Never smoker | 6,994 (61.37) |
| Former smoker | 1,754 (15.39) |
| Current smoker | 2,649 (23.24) |
| **Moderate physical activity, times/week** |  |
| 0 | 6,509 (57.11) |
| 1-2 | 1,257 (11.03) |
| 3-5 | 2,009 (17.63) |
| 6-7 | 1,622 (14.23) |
| **Depression symptoms** |  |
| No | 10,215 (89.63) |
| Yes | 1,182 (10.37) |
| **Experienced Unmet medical needs** |  |
| No | 9,052 (79.42) |
| Yes | 2,345 (20.58) |

Abbreviation: NHIS, national health insurance service.

### 2.3. Statistical analysis

LCA is a person-centered mixture modeling approach that identifies latent subpopulations within a sample based on patterns of responses to observed categorical variables ^15,38^. To determine the appropriate number of latent classes, we increased the number of classes from 2 to 5 and made decisions based on model fit statistics and theoretical interpretability **(Table S1)**. Statistical model fit was assessed using the Entropy, Akaike information criterion (AIC), Bayesian information criterion (BIC), and likelihood-ratio G2 statistic ^39^. The entropy value ranges from 0 to 1, with higher values indicating better classification accuracy. AIC and BIC are goodness-of-fit indicators to find the optimal balance between model fit and parsimony, and the G² statistic can be utilized for assessing model fit ^40^. The AIC, BIC, and G² statistics indicate that lower values reflect better-fitting models. The three-class model appears to be the best fit, as AIC and BIC show a significant decrease in the three groups. Also, the G^2^=210.8 for the three-class model shows a large drop from the two-class model with G^2^=1346.7. Finally, the highest entropy value of 0.64 in the three-class model suggests good class interpretability and separation.

To handle the time-to-event outcomes and censored observations, survival analysis can be used ^41^. Our study initially performed the proportional hazards assumption test through Schoenfeld residuals. Since the assumptions were not satisfied, the accelerated failure time (AFT) model was applied as an alternative. The AFT model is a useful approach to survival analysis, and it can be formulated using specific survival functions ^42^. In our study, the Weibull distribution was identified as the best fit based on the AIC compared to the exponential distribution. The AFT model represents time ratios, which quantify the relationship between SES and CVD incidence. The AFT model is interpreted as showing a reduction (less than 1) for a deleterious covariate and an increase (greater than 1) for a protective covariate.

Based on previous studies showing significant age-sex–standardized prevalence of CVD risk in each SES stratum ^43^, we tested the interaction between sex and age in SES on CVD incidence. Many studies assume no interaction between the exposure and mediator in their effects on the outcome, but this assumption is often unrealistic ^44^. Therefore, we examined whether there were interactions between SES and each mediator on CVD.

In traditional mediation analysis within survival analysis, the change in hazard ratios when a mediator variable is included versus excluded from the model cannot be interpreted causally ^45^. However, causal mediation analysis is more general and rigorous, as it accommodates nonlinear relationships and interactions ^46^. Therefore, we considered methodologies used for causal mediation analysis in survival outcomes using the AFT model ^47^. The natural direct effect (NDE) and natural indirect effect (NIE) were calculated using the AFT model, and the proportion mediated (PM) was estimated using the following formula ^48^. All statistical analyses were performed using SAS V.9.4 (SAS Institute, Cary, North Carolina, USA).

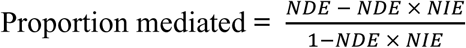

### 2.4. Ethical approval

This study was approved by the institutional review board of Seoul National University (IRB No. E2409/002-002). The requirement for informed consent was waived since the KHPS is anonymized data with no personal identifying information.

## 3. Results

### 3.1. Characteristics of the study population

The population consisted of 11,397 individuals, with men accounting for 44.98% and women accounting for 55.02% (**Table 1**). Age 60 and above accounts for the highest proportion at 31.10%. For smoking status, never-smokers represented the largest group at 61.37%, and those engaging in moderate physical activity zero times a week accounted for 57.11%, also representing the largest proportion. Participants reporting depression symptoms account for 10.37%, while 20.58% reported experiencing unmet medical needs.

### 3.2. Assessment of socioeconomic status using latent class analysis

We evaluated the characteristics of each latent class based on mean posterior and item-response probabilities (**Table 2**). The mean posterior probabilities for all latent classes exceeded the minimum threshold of 0.8, indicating an acceptable level of classification uncertainty ^38^. Based on the item-response probabilities, latent Class 1 can be defined as “medium SES” due to intermediate income and education levels. Class 2 can be characterized as “high SES” based on high income, education, and full-time working status. Class 3 can be defined as “low SES” due to low income, education levels, and no working status. Low SES accounts for 15.28% (n = 1,742), medium SES for 67.76% (n = 7,723), and high SES for 16.95% (n =1,932), with each group meeting the requirement of comprising more than 5% of the total sample. In all analyses, the reference category was set as medium SES, as it had the largest sample size and allowed for observing trends in low SES and high SES.

**Table 2.** Mean posterior probabilities and item-response probabilities in models with three latent classes.

| **Item** | **Latent class 1**<br>(n=7,723,<br>67.76%) | **Latent class 2**<br>(n=1,932,<br>16.95%) | **Latent class 3**<br>(n=1,742,<br>15.28%) |
| --- | --- | --- | --- |
| **Mean posterior probabilities** | 0.84 | 0.90 | 0.85 |
| **Item-response probabilities** |  |  |  |
| Income 1 (low: 1<sup>st</sup> quintile) | 0.05 | 0.01 | **0.79** |
| Income 2 (middle: 2<sup>nd</sup>-4<sup>th</sup> quintiles) | **0.81** | 0.41 | 0.21 |
| Income 3 (high: 5<sup>th</sup> quintiles) | 0.14 | **0.58** | 0.00 |
| Education 1 (< elementary) | 0.21 | 0.01 | **0.65** |
| Education 2 (middle-high school) | **0.60** | 0.25 | 0.29 |
| Education 3 (> college) | 0.19 | **0.74** | 0.06 |
| Working 1 (none) | **0.65** | 0.33 | **0.89** |
| Working 2 (part-time) | 0.35 | 0.24 | 0.11 |
| Working 3 (full-time) | 0.00 | **0.43** | 0.00 |
| Insurance 1 (none) | 0.00 | 0.00 | 0.00 |
| Insurance 2 (medical aid) | 0.01 | 0.00 | 0.20 |
| Insurance 3 (NHIS) | **0.99** | **1.00** | **0.80** |

Note: The maximal item-response probabilities for each latent class were marked in bold.

The distribution of SES classified according to LCA was presented in **Table 3**. Low SES constitutes the highest proportion among those aged 60 and older (total: 72.16%, men: 71.30%, women: 72.67%). Smoking exhibited different trends between men and women: among men, current smokers constituted the highest proportion across all SES groups, while among women, never smokers represented the largest percentage across all groups. In the total population, the group with the highest proportion of individuals engaging in physical activity 0 times per week was the low SES group (69.80%), while the group most frequently participating in physical activity 6–7 times per week was the medium SES group (15.87%). In the total population, the low SES group showed the highest proportion of individuals with depression (18.37%) and unmet medical needs (25.95%).

**Table 3.** Characteristics of the study population derived from latent class analysis.

| **Socioeconomic status derived from latent class analysis** |  |  |  |  |  |  |  |  |  |
| --- | --- | --- | --- | --- | --- | --- | --- | --- | --- |
| **Variables** | **Total** |  |  | **Men** |  |  | **Women** |  |  |
|  | **Low**<br>**SES** | **Medium**<br>**SES** | **High**<br>**SES** | **Low**<br>**SES** | **Medium**<br>**SES** | **High**<br>**SES** | **Low**<br>**SES** | **Medium**<br>**SES** | **High**<br>**SES** |
| **Age, N (%)** |  |  |  |  |  |  |  |  |  |
| 30-39 | 122 | 1,792 | 784 | 35 | 710 | 449 | 87 | 1,082 | 335 |
|  | (7.00) | (23.20) | (40.58) | (5.40) | (21.91) | (36.27) | (7.95) | (24.14) | (48.27) |
| 40-49 | 170 | 1,980 | 700 | 75 | 819 | 458 | 95 | 1,161 | 242 |
|  | (9.76) | (25.64) | (36.23) | (11.57) | (25.28) | (37.00) | (8.68) | (25.90) | (34.87) |
| 50-59 | 193 | 1,766 | 345 | 76 | 726 | 253 | 117 | 1,040 | 92 |
|  | (11.08) | (22.87) | (17.86) | (11.73) | (22.41) | (20.44) | (10.69) | (23.20) | (13.26) |
| 60+ | 1,257 | 2,185 | 103 | 462 | 985 | 78 | 795 | 1,200 | 25 |
|  | (72.16) | (28.29) | (5.33) | (71.30) | (30.40) | (6.30) | (72.67) | (26.77) | (3.60) |
| **Residence, N (%)** |  |  |  |  |  |  |  |  |  |
| Urban | 248 | 1,613 | 488 | 90 | 647 | 298 | 158 | 966 | 190 |
|  | (14.24) | (20.89) | (25.26) | (13.89) | (19.97) | (24.07) | (14.44) | (21.55) | (27.38) |
| Rural | 1,494 | 6,110 | 1,444 | 558 | 2,593 | 940 | 936 | 3,517 | 504 |
|  | (85.76) | (79.11) | (74.74) | (86.11) | (80.03) | (75.93) | (85.56) | (78.45) | (72.62) |
| **Marital status, N (%)** |  |  |  |  |  |  |  |  |  |
| Never married | 72<br>(4.13) | 521<br>(6.75) | 209<br>(10.82) | 45<br>(6.94) | 354<br>(10.93) | 123<br>(9.94) | 27<br>(2.47) | 167<br>(3.73) | 86<br>(12.39) |
| Married | 1,102<br>(63.26) | 6,330<br>(81.96) | 1,691<br>(87.53) | 511<br>(78.86) | 2,719<br>(83.92) | 1,094<br>(88.37) | 591<br>(54.02) | 3,611<br>(80.55) | 597<br>(86.02) |
| Separated/Divorced | 114<br>(6.54) | 259<br>(3.35) | 23<br>(1.19) | 56<br>(8.64) | 106<br>(3.27) | 18<br>(1.45) | 58<br>(5.30) | 153<br>(3.41) | 5<br>(0.72) |
| Widowed | 454<br>(26.06) | 613<br>(7.94) | 9<br>(0.47) | 36<br>(5.56) | 61<br>(1.88) | 3<br>(0.24) | 418<br>(38.21) | 552<br>(12.31) | 6<br>(0.86) |
| **Smoking, N (%)** |  |  |  |  |  |  |  |  |  |
| Never smoker | 1,095<br>(62.86) | 4,909<br>(63.56) | 990<br>(51.24) | 100<br>(15.43) | 635<br>(19.60) | 312<br>(25.20) | 995<br>(90.95) | 4,274<br>(95.34) | 678<br>(97.69) |
| Former smoker | 299<br>(17.16) | 1,081<br>(14.00) | 374<br>(19.36) | 270<br>(41.67) | 993<br>(30.65) | 361<br>(29.16) | 29<br>(2.65) | 88<br>(1.96) | 13<br>(1.87) |
| Current smoker | 348<br>(19.98) | 1,733<br>(22.44) | 568<br>(29.40) | 278<br>(42.90) | 1,612<br>(49.75) | 565<br>(45.64) | 70<br>(6.40) | 121<br>(2.70) | 3<br>(0.43) |
| **Moderate physical activity, times/week, N (%)** |  |  |  |  |  |  |  |  |  |
| 0 | 1,216<br>(69.80) | 4,355<br>(56.39) | 938<br>(48.55) | 420<br>(64.81) | 1,584<br>(48.89) | 517<br>(41.76) | 796<br>(72.76) | 2,771<br>(61.81) | 421<br>(60.66) |
| 1-2 | 85<br>(4.88) | 779<br>(10.09) | 393<br>(20.34) | 30<br>(4.63) | 392<br>(12.10) | 298<br>(24.07) | 55<br>(5.03) | 387<br>(8.63) | 95<br>(13.69) |
| 3-5 | 196 | 1,363 | 450 | 79 | 624 | 313 | 117 | 739 | 137 |
|  | (11.25) | (17.65) | (23.29) | (12.19) | (19.26) | (25.28) | (10.69) | (16.48) | (19.74) |
| 6-7 | 245 | 1,226 | 151 | 119 | 640 | 110 | 126 | 586 | 41 |
|  | (14.06) | (15.87) | (7.82) | (18.36) | (19.75) | (8.89) | (11.52) | (13.07) | (5.91) |
| **Depression symptoms, N (%)** |  |  |  |  |  |  |  |  |  |
| No | 1,422 | 6,993 | 1,800 | 558 | 2,999 | 1,166 | 864 | 3,994 | 634 |
|  | (81.63) | (90.55) | (93.17) | (86.11) | (92.56) | (94.18) | (78.98) | (89.09) | (91.35) |
| Yes | 320 | 730 | 132 | 90 | 241 | 72 | 230 | 489 | 60 |
|  | (18.37) | (9.45) | (6.83) | (13.89) | (7.44) | (5.82) | (21.02) | (10.91) | (8.65) |
| **Experienced Unmet medical needs, N (%)** |  |  |  |  |  |  |  |  |  |
| No | 1,290 | 6,189 | 1,573 | 509 | 2,679 | 1,026 | 781 | 3,510 | 547 |
|  | (74.05) | (80.14) | (81.42) | (78.55) | (82.69) | (82.88) | (71.39) | (78.30) | (78.82) |
| Yes | 452 | 1,534 | 359 | 139 | 561 | 212 | 313 | 973 | 147 |
|  | (25.95) | (19.86) | (18.58) | (21.45) | (17.31) | (17.12) | (28.61) | (21.70) | (21.18) |

Abbreviation: SES, socioeconomic status;

### 3.3. Assessment of interaction between socioeconomic status, sex, age, and mediators

We examined the interaction between SES and sex, age, and mediators. SES showed a significant interaction with sex but not with age (**Table S2**). Therefore, all subsequent analyses were stratified by sex in the AFT model. We also examined the interaction between SES and the binary mediators (**Table S3**). All mediators were converted into binary variables to use the subsequent mediator analysis using the SAS macro ^49^. When examining the interaction with the mediator by sex, a significant interaction between low SES and ever-smoking was identified in men.

### 3.4. Mediation analysis with the Accelerated failure time model

The results of the mediation analysis using the AFT model with Weibull distribution are provided in **Table 4**. The interaction results between SES and mediator variables were considered and analyzed within the model. In men, using medium SES as the reference, both low and high SES indicated a similar direction of increasing survival time trends in direct effects, but none of the results were statistically significant. In women, low SES was associated with a shorter survival time compared to medium SES for all mediators, which was statistically significant. However, high SES was associated with a longer survival time compared to medium SES, although this was not statistically significant. Natural indirect effects with CI excluding 1 were found in low SES for depression symptoms mediators in women. Low SES on average had a 21% shorter survival time until their first CVD compared to Medium SES, and 2% could be attributed to depression symptoms.

**Table 4.**
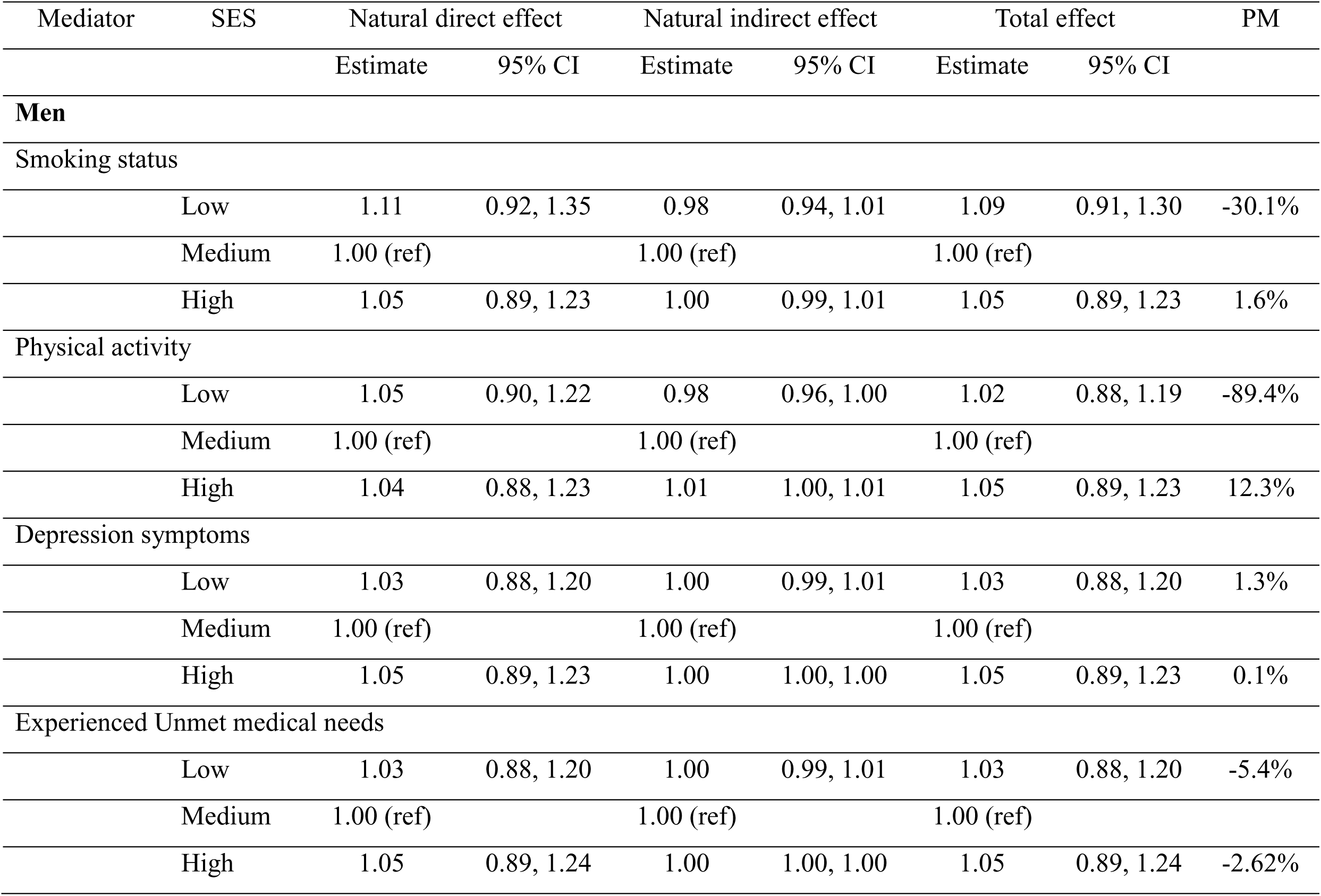
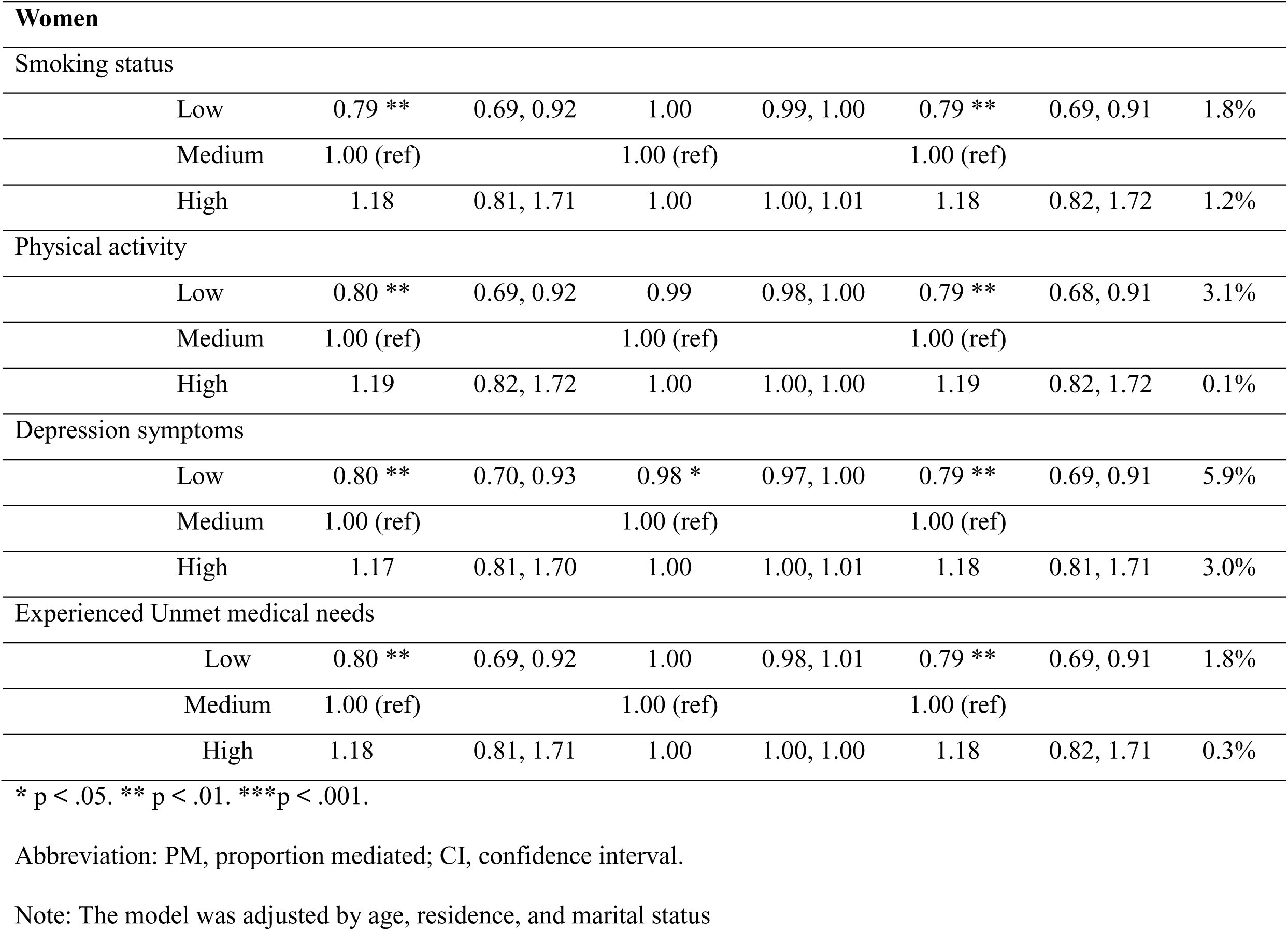
Mediation analysis with accelerated failure time models.

| Mediator | SES | Natural direct effect |  | Natural indirect effect |  | Total effect |  | PM |
| --- | --- | --- | --- | --- | --- | --- | --- | --- |
|  |  | Estimate | 95% CI | Estimate | 95% CI | Estimate | 95% CI |  |
| Men |  |  |  |  |  |  |  |  |
| Smoking status |  |  |  |  |  |  |  |  |
|  | Low | 1.11 | 0.92, 1.35 | 0.98 | 0.94, 1.01 | 1.09 | 0.91, 1.30 | -30.1% |
|  | Medium | 1.00 (ref) |  | 1.00 (ref) |  | 1.00 (ref) |  |  |
|  | High | 1.05 | 0.89, 1.23 | 1.00 | 0.99, 1.01 | 1.05 | 0.89, 1.23 | 1.6% |
| Physical activity |  |  |  |  |  |  |  |  |
|  | Low | 1.05 | 0.90, 1.22 | 0.98 | 0.96, 1.00 | 1.02 | 0.88, 1.19 | -89.4% |
|  | Medium | 1.00 (ref) |  | 1.00 (ref) |  | 1.00 (ref) |  |  |
|  | High | 1.04 | 0.88, 1.23 | 1.01 | 1.00, 1.01 | 1.05 | 0.89, 1.23 | 12.3% |
| Depression symptoms |  |  |  |  |  |  |  |  |
|  | Low | 1.03 | 0.88, 1.20 | 1.00 | 0.99, 1.01 | 1.03 | 0.88, 1.20 | 1.3% |
|  | Medium | 1.00 (ref) |  | 1.00 (ref) |  | 1.00 (ref) |  |  |
|  | High | 1.05 | 0.89, 1.23 | 1.00 | 1.00, 1.00 | 1.05 | 0.89, 1.23 | 0.1% |
| Experienced Unmet medical needs |  |  |  |  |  |  |  |  |
|  | Low | 1.03 | 0.88, 1.20 | 1.00 | 0.99, 1.01 | 1.03 | 0.88, 1.20 | -5.4% |
|  | Medium | 1.00 (ref) |  | 1.00 (ref) |  | 1.00 (ref) |  |  |
|  | High | 1.05 | 0.89, 1.24 | 1.00 | 1.00, 1.00 | 1.05 | 0.89, 1.24 | -2.62% |

| Women |  |  |  |  |  |  |  |  |
| --- | --- | --- | --- | --- | --- | --- | --- | --- |
| Smoking status |  |  |  |  |  |  |  |  |
|  | Low | 0.79 ** | 0.69, 0.92 | 1.00 | 0.99, 1.00 | 0.79 ** | 0.69, 0.91 | 1.8% |
|  | Medium | 1.00 (ref) |  | 1.00 (ref) |  | 1.00 (ref) |  |  |
|  | High | 1.18 | 0.81, 1.71 | 1.00 | 1.00, 1.01 | 1.18 | 0.82, 1.72 | 1.2% |
| Physical activity |  |  |  |  |  |  |  |  |
|  | Low | 0.80 ** | 0.69, 0.92 | 0.99 | 0.98, 1.00 | 0.79 ** | 0.68, 0.91 | 3.1% |
|  | Medium | 1.00 (ref) |  | 1.00 (ref) |  | 1.00 (ref) |  |  |
|  | High | 1.19 | 0.82, 1.72 | 1.00 | 1.00, 1.00 | 1.19 | 0.82, 1.72 | 0.1% |
| Depression symptoms |  |  |  |  |  |  |  |  |
|  | Low | 0.80 ** | 0.70, 0.93 | 0.98 * | 0.97, 1.00 | 0.79 ** | 0.69, 0.91 | 5.9% |
|  | Medium | 1.00 (ref) |  | 1.00 (ref) |  | 1.00 (ref) |  |  |
|  | High | 1.17 | 0.81, 1.70 | 1.00 | 1.00, 1.01 | 1.18 | 0.81, 1.71 | 3.0% |
| Experienced Unmet medical needs |  |  |  |  |  |  |  |  |
|  | Low | 0.80 ** | 0.69, 0.92 | 1.00 | 0.98, 1.01 | 0.79 ** | 0.69, 0.91 | 1.8% |
|  | Medium | 1.00 (ref) |  | 1.00 (ref) |  | 1.00 (ref) |  |  |
|  | High | 1.18 | 0.81, 1.71 | 1.00 | 1.00, 1.00 | 1.18 | 0.82, 1.71 | 0.3% |

\* p < .05. \*\* p < .01. \*\*\*p < .001.
Abbreviation: PM, proportion mediated; CI, confidence interval.
Note: The model was adjusted by age, residence, and marital status

## 4. Discussion

This study examined the intermediate mechanisms connecting SES and the incidence of CVD, focusing on individual health behavior, depression symptoms, and the healthcare system. We employed mediation analysis to explore the potential pathways linking SES to CVD using KHPS data, and found that depression among women exhibited a significant indirect effect.

In this study, SES was derived using four variables—family income level, education level, working status, and health insurance—through LCA, resulting in three distinct classes. This study identified a detailed SES profile that traditional SES indicators, such as education or occupation, could not capture. While income and education were categorized into three levels (low, middle, and high) for each latent SES class, many individuals with working status were included in the “none” category, except for those in high SES. This suggests that medium SES’s income may be composed of other resources rather than individual earnings. Therefore, using LCA to define SES allows for identifying underlying aspects that cannot be captured when defined solely by family income.

When stratified by sex, a significant direct effect of low SES on increased CVD risk was observed among women, while no significant effects were found among men. Our study is consistent with the findings regarding gender differences in the association between SES and cardiovascular risk, showing that lower SES was associated with higher CVD risk only for women ^50^. According to the existing systematic review, women show a higher incidence risk of CVD compared to men with low education ^51^, and in low to middle-income, women exhibit a more sensitive risk of CVD than men ^7^. As CVD incidence may differ by sex even among those with similar SES, the consideration of gender-specific strategies, especially for women with lower SES, is essential.

Several theories have been proposed to clarify why SES might be more strongly linked to CVD for women than for men. First, the differences in the association between SES and CVD by sex are related to the access and adherence to preventative treatment and CVRF management across varying levels of socioeconomic disadvantage ^51^. The delay in the time to diagnosis or treatment, resulting from a longer duration until presentation, is often attributed to women’s lower risk awareness ^52^. Second, women with lower educational attainment may be more likely than men to experience social and familial circumstances, such as single parenting, that contribute to elevated cardiovascular risk ^53^. Third, women with lower SES tend to experience menopause earlier, which may alter body composition and increase the duration of exposure to heightened CVD risk ^54,55^.

The AFT model showed significant indirect effects for the depression symptoms in women, indicating that the presence of depressive symptoms decreased the average survival times from CVD and mediated 5.9%. Several studies suggest that women with low SES were found to have a higher likelihood of experiencing a major depressive episode ^56,57^. Other studies suggest that sex interacts with household income on major depressive episodes, indicating that the association between them is larger for women ^58^. Women are more vulnerable to the effects of environmental triggers for depression, such as low SES, than men^59^. Psychosocial gender role-related risk factors may contribute to the higher risk of depression in women, reflecting sex differences in endocrine stress reactions ^60–62^.

Depression is a risk factor for CVD, and its impact on CVD risk is greater in women than in men ^63,64^. The mechanisms explaining the relationship between depression and CVD in women can be attributed to low-grade chronic inflammation, accompanied by increases in inflammatory markers, platelet activation, coagulation, and heightened sympathetic tone ^65^. Additionally, in women, changes in sex hormones over their lifetimes have a significant impact on depression and cardiovascular health ^65^. Sex hormones such as estrogen and progesterone inhibit pro-inflammatory cytokines and increase the production of anti-inflammatory cytokines, which are more directly related to inflammation patterns ^66^. Combined with CVRFs and reproductive hormone fluctuations, it may exacerbate the inflammatory response, contributing to the depression-CVD relationship ^65,67^.

This study focused on exploring the pathways linking SDoH to CVD, not only directly, but also indirectly through midstream risk factors (psychological stress, physical inactivity, and smoking behaviors) ^23^. In women, some indirect intervention needs were identified in depression, but the effects were minimal. This suggests that interventions targeting upstream determinants, such as addressing the basic socioeconomic distribution and financial barriers due to unfavorable social conditions, are also important. It aligns with the American Heart Association’s recommendation to broaden SDoH interventions to address upstream determinants of CVD, such as poverty and education ^68^.

Our study has several strengths. First, we applied overall SES using LCA, providing a detailed SES profile that each traditional SES indicator could not capture. Including various SES-related factors to derive latent classes helps mitigate potential biases in causal direction that may arise when used alone. Second, quantitative assessment of mediator effects can prioritize risk factors to address such health inequalities. We employed a counterfactual causal mediation approach, which offers robust causal insights and can accommodate potential interactions between exposure and mediator. The AFT model is preferred over the

Cox model in mediation analysis, as it does not require the rare outcome assumption ^49,69^.

Third, longitudinal study design offers better insights into the causal relationship between SES, mediators, and CVD incidence than cross-sectional studies.

Despite the strengths noted above, our studies have several limitations. First, to establish the validity of the SES derived through LCA, it is necessary to compare it with other established composite SES indicators. Second, health behavior, depression symptoms, and unmet medical needs were self-reported, which can lead to misclassification bias due to memory loss. Third, in the AFT models, the mediator variable was dichotomized for the analysis, which did not fully capture the information available in the original mediator variable’s categories. Fourth, the CVD incidence was 8.8% among all study populations, indicating that there were few cases in the study, which resulted in limited statistical power to detect the effect. Fifth, the assumptions for causal mediation analysis, including no mediator-outcome confounding affected by exposure variables, and no unmeasured confounding, can be violated in our studies. For example, the mediator-outcome confounding assumption does not hold with smoking status or physical activity. Depression can affect smoking status or physical activity and CVD, and it can be affected by SES ^70–72^. Also, genetic factors, social support, and environmental factors could be considered as unmeasured confounders that could bias the estimates of SES-mediator variables and mediator-CVD relationships. Lastly, mediation analysis can only demonstrate causal effects if confounding can be ruled out. If multiple mediators are involved, there is a potential for post-randomization confounders affecting the effects of other mediators, requiring caution in interpretation ^73^. Our study uses one model for each mediator, but further studies using a multiple-mediator model through different causal pathways should be considered.

## 5. Conclusion

This study examined the relationship between SES profiles and CVD incidence and explored the indirect effects of health behavior, depression, and unmet medical needs. We observed a sex difference in the association between SES and CVD. The causal mediation analysis revealed the indirect pathways through which low and high SES influenced CVD incidence, providing important insights into the mechanisms driving socioeconomic inequalities in CVD risk. Although the effect is modest, depression symptoms among women contribute to SES disparities in CVD incidence. This suggests that the risk of CVD can be attributed to the differential distributions of depression symptoms across SES subgroups. Policies should be considered to intervene in the pathways from low SES to depression symptoms in women to prevent CVD risk. However, given the small proportion of the mediated effect, interventions at the upstream levels to address SES inequalities and their impact on health outcomes are essential.

## Data Availability

The raw data are accessible to everyone and can be obtained by submitting a requisition form through the official KHPS website (www.khp.re.kr:444/).

www.khp.re.kr:444/

## Acknowledgments Source of Funding

None.

## Supplementary Material

Tables S1-S3

## Non-standard Abbreviations and Acronyms

AFT: accelerated failure time
AIC: Akaike information criterion
BIC: Bayesian information criterion
CI: confidence interval
CSDH: Commission on Social Determinants of Health
CVD: Cardiovascular disease
CVRFs: cardiovascular risk factors
DALYs: disability-adjusted life-year
ICD-10: International Classification of Diseases 10th edition
KHPS: Korea Health Panel Survey
KIHASA: Korea Institute for Health and Social Affairs
LCA: latent class analysis
NDE: natural direct effect
NHIS: National Health Insurance Service
NIE: natural indirect effect
SDoH: social determinants of health
SES: socioeconomic status
YLLs: years of life lost

## References

1. Tsao CW, Aday AW, Almarzooq ZI, Anderson CA, Arora P, Avery CL, Baker-Smith CM, Beaton AZ, Boehme AK, Buxton AE. Heart disease and stroke statistics—2023 update: a report from the American Heart Association. Circulation. 2023;147:e93–e621.

2. Kim HC. Epidemiology of cardiovascular disease and its risk factors in Korea. Global health & medicine. 2021;3:134–141.

3. Shin JI, Oh J, Kim HC, Choi D, Yoon Y-s. Current state of cardiovascular research in Korea. In: Am Heart Assoc; 2019.

4. Dahlöf B. Cardiovascular disease risk factors: epidemiology and risk assessment. The American journal of cardiology. 2010;105:3A–9A.

5. Kozakiewicz K, Podolecka E, Kwaśniewska M, Drygas W, Pająk A, Tendera M. Association between socioeconomic status and cardiovascular risk. Polish Heart Journal (Kardiologia Polska). 2016;74:179–184.

6. Pourfarzi F, Moghadam TZ, Zandian H. Decomposition of Socioeconomic Inequality in Cardiovascular Disease Prevalence in the Adult Population: A Cohort-based Cross-sectional Study in Northwest Iran. Journal of Preventive Medicine and Public Health. 2022;55:297.

7. Wang T, Li Y, Zheng X. Association of socioeconomic status with cardiovascular disease and cardiovascular risk factors: a systematic review and meta-analysis. Journal of Public Health. 2024;32:385–399.

8. Winkleby MA, Jatulis DE, Frank E, Fortmann SP. Socioeconomic status and health: how education, income, and occupation contribute to risk factors for cardiovascular disease. American journal of public health. 1992;82:816–820.

9. Schultz WM, Kelli HM, Lisko JC, Varghese T, Shen J, Sandesara P, Quyyumi AA, Taylor HA, Gulati M, Harold JG. Socioeconomic status and cardiovascular outcomes: challenges and interventions. Circulation. 2018;137:2166–2178.

10. Adler N, Bush NR, Pantell MS. Rigor, vigor, and the study of health disparities. Proceedings of the National Academy of Sciences. 2012;109:17154–17159.

11. Braveman PA, Cubbin C, Egerter S, Chideya S, Marchi KS, Metzler M, Posner S. Socioeconomic status in health research: one size does not fit all. Jama. 2005;294:2879–2888.

12. Cabrera JC, Karl SR, Rodriguez MC, Chavez C. Investigating socioeconomic status proxies: is one Proxy Enough? 2018.

13. PT C. Measuring socioeconomic status: reliability and preliminary validity for different approaches. Assessment. 2002;9:145–155.

14. Lee C, Yi J-S. Socioeconomic classes among oldest-old women in South Korea: a latent class analysis. International Journal of Environmental Research and Public Health. 2021;18:13183.

15. Vermunt JK, Magidson J. Latent class analysis. The sage encyclopedia of social sciences research methods. 2004;2:549–553.

16. Hagenaars JA, McCutcheon AL. Applied latent class analysis. Cambridge University Press; 2002.

17. Zhang Y-B, Chen C, Pan X-F, Guo J, Li Y, Franco OH, Liu G, Pan A. Associations of healthy lifestyle and socioeconomic status with mortality and incident cardiovascular disease: two prospective cohort studies. Bmj. 2021;373.

18. Fairley L, Cabieses B, Small N, Petherick ES, Lawlor DA, Pickett KE, Wright J. Using latent class analysis to develop a model of the relationship between socioeconomic position and ethnicity: cross-sectional analyses from a multi-ethnic birth cohort study. BMC public health. 2014;14:1–14.

19. Lowthian E, Page N, Melendez-Torres G, Murphy S, Hewitt G, Moore G. Using latent class analysis to explore complex associations between socioeconomic status and adolescent health and well-being. Journal of Adolescent Health. 2021;69:774–781.

20. Powell-Wiley TM, Baumer Y, Baah FO, Baez AS, Farmer N, Mahlobo CT, Pita MA, Potharaju KA, Tamura K, Wallen GR. Social determinants of cardiovascular disease. Circulation research. 2022;130:782–799.

21. Solar O, Irwin A. A conceptual framework for action on the social determinants of health. In: WHO Document Production Services; 2010.

22. Hafeman DM. A sufficient cause based approach to the assessment of mediation. European journal of epidemiology. 2008;23:711–721.

23. Jilani MH, Javed Z, Yahya T, Valero-Elizondo J, Khan SU, Kash B, Blankstein R, Virani SS, Blaha MJ, Dubey P. Social determinants of health and cardiovascular disease: current state and future directions towards healthcare equity. Current atherosclerosis reports. 2021;23:1–11.

24. Dhar AK, Barton DA. Depression and the link with cardiovascular disease. Frontiers in psychiatry. 2016;7:33.

25. Pampel FC, Krueger PM, Denney JT. Socioeconomic disparities in health behaviors. Annual review of sociology. 2010;36:349–370.

26. Miller GE, Chen E, Shimbo D. Mechanistic understanding of socioeconomic disparities in cardiovascular disease. In: American College of Cardiology Foundation Washington, DC; 2019:3256-3258.

27. Hill HD, Ybarra MA. Less-Educated Workers’ Unstable Employment: Can the Safety Net Help? Fast Focus. No. 19-2014. Institute for Research on Poverty. 2014.

28. Holahan J. The 2007–09 recession and health insurance coverage. Health Affairs. 2011;30:145–152.

29. Udupa K, Sathyaprabha T, Thirthalli J, Kishore K, Lavekar G, Raju T, Gangadhar B. Alteration of cardiac autonomic functions in patients with major depression: a study using heart rate variability measures. Journal of affective disorders. 2007;100:137–141.

30. Kim Y, Kim S, Jeong S, Cho SG, Hwang S-s. Poor people and poor health: examining the mediating effect of unmet healthcare needs in Korea. Journal of Preventive Medicine and Public Health. 2019;52:51.

31. Kim B-R, Cho H-A, Shin H. The effects of orthodontic treatment on personal dental expenditures in South Korea: a follow-up study using Korean health panel survey. BMC Health Services Research. 2022;22:1598.

32. Benjamin EJ, Muntner P, Alonso A, Bittencourt MS, Callaway CW, Carson AP, Chamberlain AM, Chang AR, Cheng S, Das SR. Heart disease and stroke statistics—2019 update: a report from the American Heart Association. Circulation. 2019;139:e56–e528.

33. Son JS, Choi S, Kim K, Kim SM, Choi D, Lee G, Jeong S-M, Park SY, Kim Y-Y, Yun J-M. Association of blood pressure classification in Korean young adults according to the 2017 American College of Cardiology/American Heart Association guidelines with subsequent cardiovascular disease events. Jama. 2018;320:1783–1792.

34. Lee G, Jeong S, Choi S, Kim KH, Chang J, Kim SR, Kim K, Son JS, Kim SM, Choi D. Associations between alcohol consumption and cardiovascular disease among long-term survivors of colorectal cancer: a population-based, retrospective cohort study. BMC cancer. 2021;21:710.

35. Zhu Y, Llamosas-Falcón L, Kerr WC, Rehm J, Probst C. Behavioral risk factors and socioeconomic inequalities in ischemic heart disease mortality in the United States: A causal mediation analysis using record linkage data. PLoS medicine. 2024;21:e1004455.

36. Nordahl H, Rod NH, Frederiksen BL, Andersen I, Lange T, Diderichsen F, Prescott E, Overvad K, Osler M. Education and risk of coronary heart disease: assessment of mediation by behavioral risk factors using the additive hazards model. European journal of epidemiology. 2013;28:149–157.

37. Hua J, Shen R, Guo X, Yu L, Qiu M, Ma L, Peng X. The mediating role of depression in the association between socioeconomic status and cardiovascular disease: A nationwide cross-sectional study from NHANES 2005–2018. Journal of Affective Disorders. 2024;366:466–473.

38. Weller BE, Bowen NK, Faubert SJ. Latent class analysis: a guide to best practice. Journal of black psychology. 2020;46:287–311.

39. Lanza ST, Collins LM, Lemmon DR, Schafer JL. PROC LCA: A SAS procedure for latent class analysis. Structural equation modeling: a multidisciplinary journal. 2007;14:671–694.

40. Collins LM, Fidler PL, Wugalter SE, Long JD. Goodness-of-fit testing for latent class models. Multivariate Behavioral Research. 1993;28:375–389.

41. George B, Seals S, Aban I. Survival analysis and regression models. Journal of nuclear cardiology. 2014;21:686–694.

42. Martinussen T, Peng L. Alternatives to the Cox model. Handook of Survival Analysis. 2013.

43. Kanjilal S, Gregg EW, Cheng YJ, Zhang P, Nelson DE, Mensah G, Beckles GL. Socioeconomic status and trends in disparities in 4 major risk factors for cardiovascular disease among US adults, 1971-2002. Archives of internal medicine. 2006;166:2348-2355.

44. Valeri L, VanderWeele TJ. Mediation analysis allowing for exposure–mediator interactions and causal interpretation: theoretical assumptions and implementation with SAS and SPSS macros. Psychological methods. 2013;18:137.

45. Lange T, Hansen JV. Direct and indirect effects in a survival context. Epidemiology. 2011;22:575–581.

46. Lee H, Herbert RD, McAuley JH. Mediation analysis. Jama. 2019;321:697–698.

47. VanderWeele TJ. Causal mediation analysis with survival data. Epidemiology. 2011;22:582–585.

48. VanderWeele TJ, Vansteelandt S. Odds ratios for mediation analysis for a dichotomous outcome. American journal of epidemiology. 2010;172:1339–1348.

49. Valeri L, VanderWeele TJ. SAS macro for causal mediation analysis with survival data. Epidemiology. 2015;26:e23–e24.

50. Jenkins KR, Ofstedal MB. The association between socioeconomic status and cardiovascular risk factors among middle-aged and older men and women. Women & health. 2014;54:15–34.

51. Backholer K, Peters SA, Bots SH, Peeters A, Huxley RR, Woodward M. Sex differences in the relationship between socioeconomic status and cardiovascular disease: a systematic review and meta-analysis. J Epidemiol Community Health. 2017;71:550–557.

52. Mehta LS, Beckie TM, DeVon HA, Grines CL, Krumholz HM, Johnson MN, Lindley KJ, Vaccarino V, Wang TY, Watson KE. Acute myocardial infarction in women: a scientific statement from the American Heart Association. Circulation. 2016;133:916–947.

53. Loucks EB, Sullivan LM, Hayes LJ, D’Agostino Sr RB, Larson MG, Vasan RS, Benjamin EJ, Berkman LF. Association of educational level with inflammatory markers in the Framingham Offspring Study. American journal of epidemiology. 2006;163:622–628.

54. Vélez MP, Alvarado BE, Lord C, Zunzunegui M-V. Life course socioeconomic adversity and age at natural menopause in women from Latin America and the Caribbean. Menopause. 2010;17:552–559.

55. Ghosh A, Bhagat M. Anthropometric and body composition characteristics in pre-and postmenopausal Asian Indian women: Santiniketan women study. Anthropologischer Anzeiger. 2010:1–10.

56. Wang JL, Schmitz N, Dewa C. Socioeconomic status and the risk of major depression: the Canadian National Population Health Survey. Journal of Epidemiology & Community Health. 2010;64:447–452.

57. Back JH, Lee Y. Gender differences in the association between socioeconomic status (SES) and depressive symptoms in older adults. Archives of gerontology and geriatrics. 2011;52:e140–e144.

58. Assari S. Social determinants of depression: The intersections of race, gender, and socioeconomic status. Brain sciences. 2017;7:156.

59. Radloff LS, Rae DS. Components of the sex difference in depression. Research in community & mental health. 1981.

60. Kuehner C. Gender differences in unipolar depression: an update of epidemiological findings and possible explanations. Acta Psychiatrica Scandinavica. 2003;108:163–174.

61. Möller-Leimkühler AM. Gender differences in cardiovascular disease and comorbid depression. Dialogues in clinical neuroscience. 2007;9:71–83.

62. Kessler RC, McGonagle KA, Swartz M, Blazer DG, Nelson CB. Sex and depression in the National Comorbidity Survey I: Lifetime prevalence, chronicity and recurrence. Journal of affective disorders. 1993;29:85–96.

63. Humphries KH, Izadnegahdar M, Sedlak T, Saw J, Johnston N, Schenck-Gustafsson K, Shah R, Regitz-Zagrosek V, Grewal J, Vaccarino V. Sex differences in cardiovascular disease– impact on care and outcomes. Frontiers in neuroendocrinology. 2017;46:46–70.

64. Lazzarino AI, Hamer M, Stamatakis E, Steptoe A. The combined association of psychological distress and socioeconomic status with all-cause mortality: a national cohort study. JAMA internal medicine. 2013;173:22–27.

65. Rivera MAM, Rivera IR, Avila W, Marques-Santos C, Costa FA, Ferro CR, Fernandes JMG. Depression and cardiovascular disease in women. International Journal of Cardiovascular Sciences. 2022;35:537–545.

66. Mattina GF, Van Lieshout RJ, Steiner M. Inflammation, depression and cardiovascular disease in women: the role of the immune system across critical reproductive events. Therapeutic advances in cardiovascular disease. 2019;13:1753944719851950.

67. Shao M, Lin X, Jiang D, Tian H, Xu Y, Wang L, Ji F, Zhou C, Song X, Zhuo C. Depression and cardiovascular disease: Shared molecular mechanisms and clinical implications. Psychiatry research. 2020;285:112802.

68. White-Williams C, Rossi LP, Bittner VA, Driscoll A, Durant RW, Granger BB, Graven LJ, Kitko L, Newlin K, Shirey M. Addressing social determinants of health in the care of patients with heart failure: a scientific statement from the American Heart Association. Circulation. 2020;141:e841–e863.

69. Gelfand LA, MacKinnon DP, DeRubeis RJ, Baraldi AN. Mediation analysis with survival outcomes: accelerated failure time vs. proportional hazards models. Frontiers in psychology. 2016;7:423.

70. Fluharty M, Taylor AE, Grabski M, Munafò MR. The association of cigarette smoking with depression and anxiety: a systematic review. Nicotine & Tobacco Research. 2016;19:3–13.

71. Roshanaei-Moghaddam B, Katon WJ, Russo J. The longitudinal effects of depression on physical activity. General hospital psychiatry. 2009;31:306–315.

72. Everson SA, Maty SC, Lynch JW, Kaplan GA. Epidemiologic evidence for the relation between socioeconomic status and depression, obesity, and diabetes. Journal of psychosomatic research. 2002;53:891–895.

73. VanderWeele T, Vansteelandt S. Mediation analysis with multiple mediators. Epidemiologic methods. 2014;2:95–115.

